# Increased vulnerability to SARS-CoV-2 infection among indigenous peoples living in the urban area of Manaus

**DOI:** 10.1101/2021.04.05.21254937

**Authors:** Gemilson Soares Pontes, Jean de Melo Silva, Renato Pinheiro-Silva, Anderson Nogueira Barbosa, Luciano Cardenes Santos, Antônio de Pádua Quirino Ramalho, Carlos Eduardo de Castro Alves, Danielle Furtado da Silva, Leonardo Calheiros de Oliveira, Allyson Guimarães da Costa, Ana Carla Bruno

## Abstract

**Background:** The COVID-19 pandemic threatens indigenous peoples living in suburban areas of large Brazilian cities and has thus far intensified their pre-existing socio-economic inequalities. This study evaluated the epidemiological situation of SARS-CoV-2 infection among residents of the biggest urban multiethnic indigenous community of the Amazonas state, Brazil.

**Methods:** Blood samples of 280 indigenous people who live in the urban community known as Parque das Tribos, which is located in the surrounding area of Manaus, were tested for the presence of anti- SARS-CoV-2 IgA or IgG antibodies using an enzyme-linked immunosorbent assay. An epidemiological standardized interviewer-administered questionnaire was applied to assess the risk factors and sociodemographic information of the study population.

**Results:** We found a total positivity rate of 64.64% (95% CI 59.01-70.28) for SARS-CoV-2 infection. IgA and IgG were detected in 55.71% (95% CI 49.89-61.54) and 60.71% (95% CI 54.98-66.45) of the individuals tested, respectively. From the total number (n=280), 80.11% of positive individuals (95%; CI 74.24-85.98) were positive for both IgA and IgG Abs. All individuals with COVID-19-related symptoms on the day of blood collection (n=11) were positive for IgG, while IgA was detected in 84.61% (n=55) of individuals who had presented symptoms several weeks before the blood collection. Individuals aged 30-39 were more susceptible to SARS-CoV-2 infection (prevalence ratio [PR] 0.77; 95% CI 0.58-1.03; p=0.033). People whose main source of information on COVID-19 was religious leaders or friends showed higher susceptibility to infection (PR 1.22; 95% CI 1.00-1.49; p=0.040). In addition, individuals who left home more frequently were at higher risk of infection (PR 1.22; 95% CI 1.00-1.49; p=0.048). Five or more individuals per household increased almost 5-fold the risk of virus transmission (Odds ratio [OR] 2.56; 95% CI; 1.09-6.01; p=0.019). Over 95% of the study population had no access to clean water and/or sanitation.

**Conclusions:** The disproportionate dissemination of SARS-CoV-2 infection observed in the Parque das Tribos urban indigenous community might be driven by typical cultural behavior and socioeconomic inequalities. Despite the pandemic threat, this population is not being targeted by public policies and appears to be chronically invisible to the Brazilian authorities.

## 1. Introduction

The state of Amazonas in northern Brazil is one of the states that has been hit hardest by the current pandemic. In the period between the WHO declaring the COVID-19 pandemic on March 11, 2020 and the end of March 2021 (1), the death toll from COVID-19 in the Amazonas state has surpassed the twelve thousand mark. The Amazonas state is located in a region of paramount environmental and demographic importance. The state comprises one-fifth of the Brazilian territory and contains approximately 40% of the world’s rainforests (2). The majority of the population in the state has an indigenous mixed-heritage background, and sixty percent of its 4 four million inhabitants live in the capital Manaus (3).

Based on the last population census, around 168,000 indigenous peoples distributed in 641 communities and 62 ethnic groups live within Amazonas state (4). Indigenous populations have to deal with many social inequalities resulting from socio-economic marginalization, including poor nutrition, lack of access to health services and proper sanitation (5). This situation put indigenous communities under worrying threat during public health emergencies, such as the coronavirus pandemic.

The number of indigenous people affected by COVID-19 in Brazil is not accurately notified. According to the Special Secretariat of Indigenous Health (SESAI), over 600 individuals died from COVID-19 in Brazil and the state of Amazonas is responsible for 25% of this number(6). However, the death rates must be much higher than the official numbers. Also the epidemiological data is not stratified by ethnicity, which could allow identifying related risk factors. On top of this, Brazil’s indigenous agencies do not monitor the COVID-19 epidemiological situation in urban indigenous communities.

There are approximately 315,000 indigenous people grouped in 300 ethnicities living close to the large cities of Brazil (4). However, the true number of indigenous populations residing in the surrounding areas of Manaus is unknown, though, it is estimated that 20,000 individuals from 92 ethnicities live in 62 urban indigenous communities (7). This population is completely invisible to the Brazilian public health agenda, especially regarding COVID-19. Thus, this study assessed the magnitude of the spread of SARS-CoV-2 and its transmission risk factors among the biggest multiethnic urban indigenous community located on the west side of Manaus, Amazonas state, Brazil.

## 2. Material and Methods

### 2.1 Ethical approval

This study was approved by the Brazilian Commission for Ethics in Research - CONEP (approval number: 4.260.763). The confidentiality and the right to leave the study at any time were guaranteed to all participants. All individuals signed the informed consent form before taking part in this study.

### 2.2 Study design and Population

Between October 10 and 14, 2020, we randomly recruited indigenous residents in the community of Parque das Tribos, the biggest multiethnic urban indigenous community in Manaus. Over 1,300 indigenous people from 35 different ethnicities live in this community, which is located in the Tarumã district, west Manaus, Amazonas state, Brazil.

From a total of 280 individuals of both genders aged from 1 to 83 years, and from different ethnicities, approximately 5 ml of peripheral blood was collected by venipuncture for posterior serological analysis. A standardized interviewer-administered questionnaire was used to obtain information on sociodemographic and risk factor variables. Because of the questionnaire’s complexity, it was decided by a multidisciplinary team (composed of anthropologists, physicians, virologists and epidemiologists) that only individuals over 13 years old would answer the questionnaire in order to guarantee the quality of the results obtained in the survey (8). Some adults (n=43) agreed to have blood collected, but refused to respond the questionnaire due to cultural issues or due to feeling uncomfortable doing so (Figure 1).

**Figure 1.**
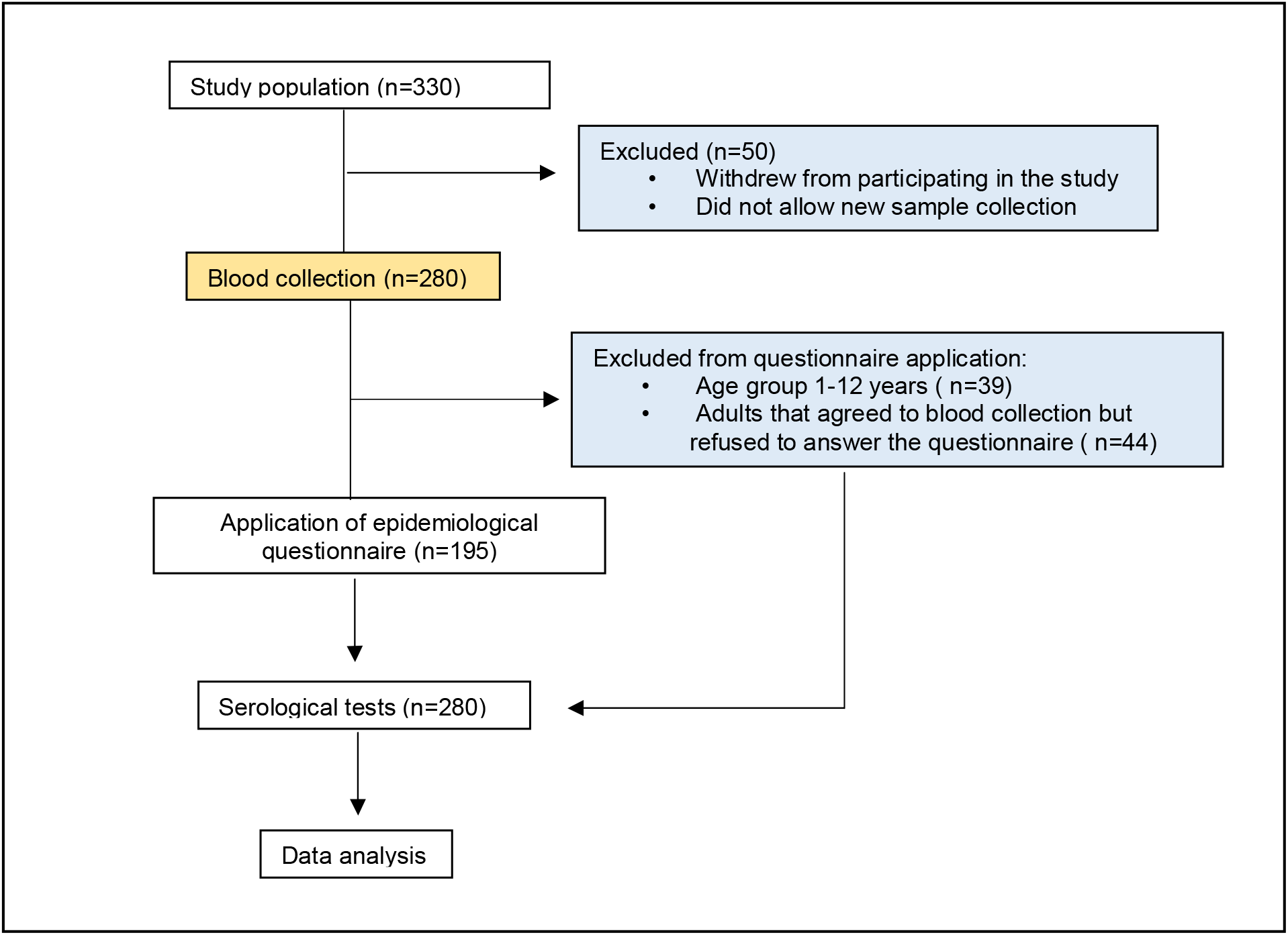
Flow chart shows study design and the number of individuals included.

### 2.3 Serological analysis

Serum samples from the study population were tested for SARS-CoV-2 IgA and IgG antibodies (Abs) through an enzyme-linked immunosorbent assay (*Anti-SARS-CoV-2 IgA/IgG ELISA-euroimmun, Germany)*, which was performed according to the manufacturer’s instructions. The optical density was measured in a spectrophotometer (Epoch Microplate, Biotek Instruments Inc. USA) using a 450 nm filter, and the test positivity was determined according to the cut-off formula indicated by the manufacturer. Cut-off ranges were obtained by calculating a ratio of the optical density (OD) values of the control or patient sample over the OD of the calibrator, according to the following formula: OD of the control or patient sample/OD of calibrator = ratio. Ratios <0.8 were considered negative and ratios > 1.1 positive. Ratios 0.8 ≥ to ≤ 1.1 were considered indeterminate. The serological tests were performed at the Laboratory of Virology and Immunology of the National Institute of Amazonian Research (INPA).

### 2.4 Statistical Analysis

Descriptive statistical analysis was used to evaluate the sociodemographic variables and serum levels of SARS-Cov-2 antibodies observed in the study population. The prevalence ratio (PR) analysis, using chi-square statistic test with Yates correction was performed to assess the relationship between sociodemographic characteristics and positivity to SARS-CoV-2 infection. The prevalence ratio analysis was performed on the following variables: genders, age, marital status, level of schooling, housing type, COVID-19 information sources and frequency of leaving the home. One-way ANOVA and a Tukey post hoc test were used to compare serum levels of SARS-CoV-2 antibodies (optical density values) among age ranges. The effect of household crowding on the risk of exposure to infection was evaluated through odds ratio (OR) analysis and Fisher’s exact test. All descriptive statistical analyses in graph format, ANOVA and Tukey’s test were performed using the R program v.4.0.4, while the prevalence ratio (PR) was performed using the Toolkit program v.3.0.1. In all analyses, a value of p < 0.05 was considered significant. A flowchart showing the steps taken in the study is shown below (Figure 1).

## 3. Results

### 3.1. Positivity rates of SARS-CoV-2 infection

The study population was composed of 28 different indigenous ethnic groups. Baré, Tukano, Tikuna and Kokama were the most common ethnicities (Figure 2A). A total of 181 individuals (64.64%; 95% CI 59.01-70.28) were positive for SARS-CoV-2 specific IgA or IgG antibodies (Abs). The positivity rates for IgA and IgG observed in the study population were 60.71% (95% CI 54.98-66.45) and 55.71% (95% CI 49.89-61.54), respectively. From the total number of individuals positive for SARS-CoV-2, 145 (80.11%;95% CI 74.24-85.98) tested positive for both IgA and IgG, whereas 11 (6.07%;95% CI 2.56-9.59) and 25 (13.81%; 95% CI 8.72-18.85) individuals were positive only for IgA or IgG, respectively (Figure 2B).

**Figure 2.**
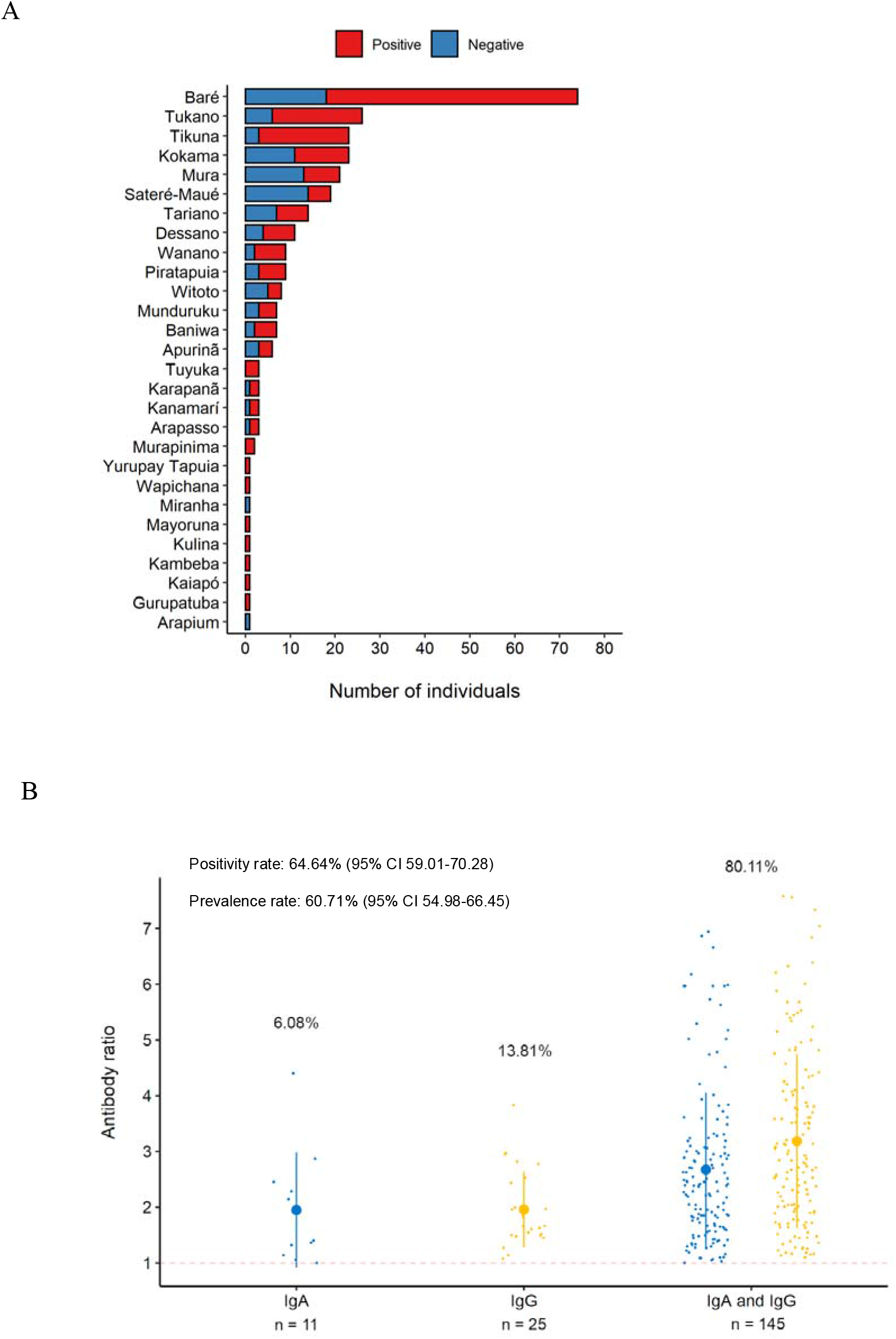
Ethnic diversity of the study population and positivity rates. (A) SARS-CoV-2 positivity according to ethnicity. (B) Pattern of IgA, IgG and IgA/IgG positivity among infected individuals. Smaller dots represent the antibody ratio from each individual. Larger dots and vertical lines represent mean and standard deviation, respectively. Dashed line represent cut-off values specified by the manufacturer.

Mild or moderate COVID-19-related symptoms were present in 107 individuals (59.11%) who were positive for SARS-CoV-2. The most common symptoms were headache, fever, body aches, loss of taste or smell and/or joint pain (Figure 3A). All individuals with symptoms on the day of blood collection (n=11) were positive for both IgA and IgG, while 88.71% (n=55) of the individuals were still displaying high serum levels of IgA even 4 weeks after the onset of their symptoms (Figure 3B).

**Figure 3.**
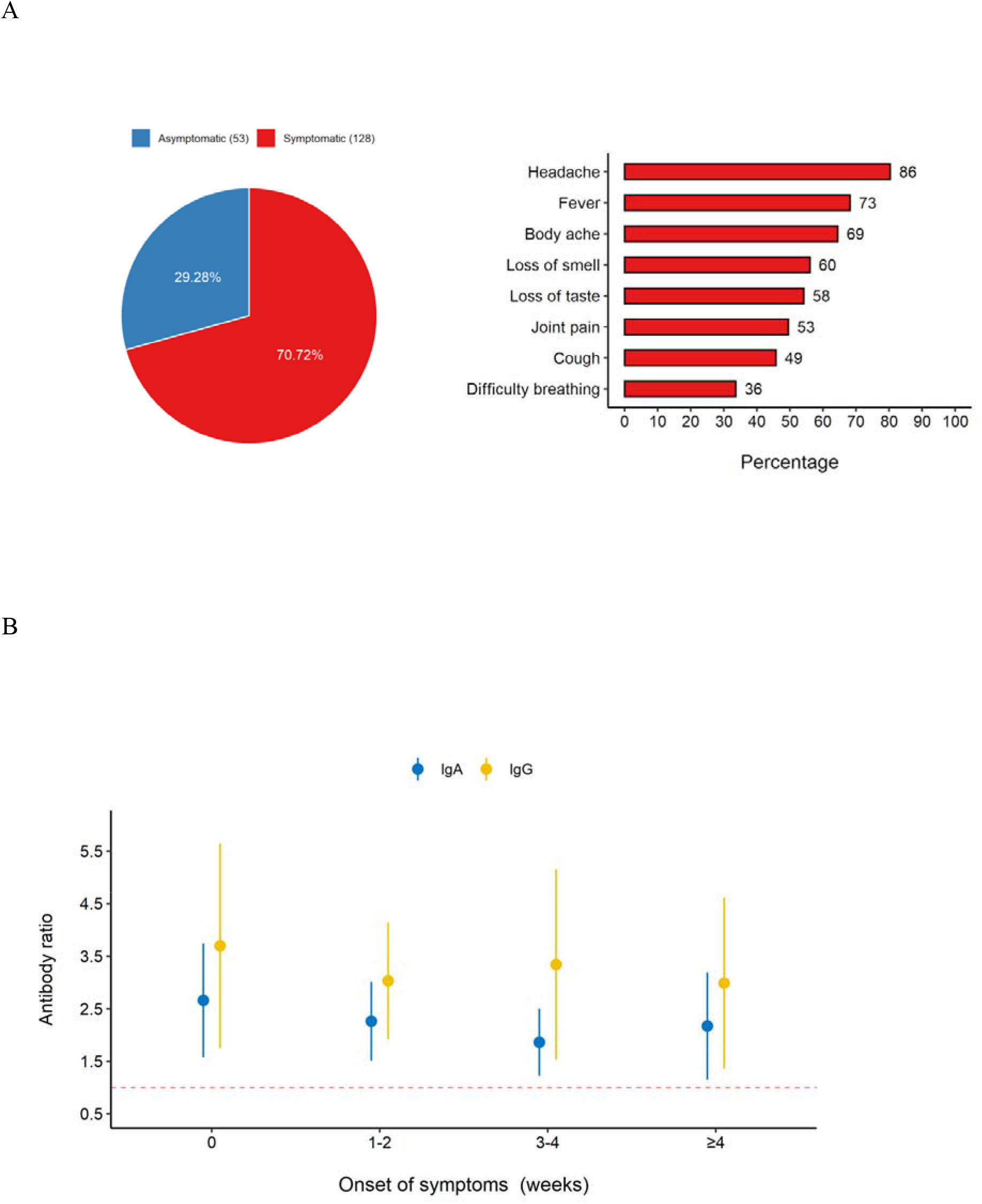
Main clinical symptoms and pattern of positivity according to the day of onset of the individual’s symptoms. (A) Number of symptomatic and asymptomatic individuals positive for SARS-CoV-2. (B) IgA and IgG positivity rates according to the day of the onset of the symptoms. Number 0 means the individuals presented symptoms on the day of blood collection. Dots and vertical lines represent mean and standard deviation, respectively. Dashed line represent cut-off values specified by the manufacturer.

### 3.2 Serological profile according to the social demographical characteristics

The positivity rates were similar between women and men (p=0.207) and the age range 30-39 years was found to be the most susceptible to SARS-CoV-2 infection (95% CI: 0.58-1.03; p=0.033) (Table 1). However, when the IgG positivity rates were compared based on antibody ratios, the difference observed between the following age groups was statistically significant: ages 20-29 and 40-49 (p=0.044); 13-19 and ≥60 (p=0.002); 20-29 and ≥60 (p=0.0004) (Supplementary 1A). People belonging to the age group ≥60 years old showed the highest antibody ratios (IgA mean ratio=3.080 ± 1.623; IgG mean ratio=4.221 ± 1.832), while the age groups 13-19 and 20-29 showed the lowest IgA (mean ratio=2.268 ± 0.919) and IgG ratios (mean ratio= 2.207 ± 1.246), respectively. These values may indicate the serum level of antibodies in these individuals, however, in order to confirm this information it is necessary to perform antibody titration. Due to budget limitations, we were not able to assess the antibody serum levels.

**Table 1.** SARS-CoV-2 positivity according to sociodemographic characteristics of the study population

| Sociodemographic Characteristics | N (%) | Age Range | IgA N (%) | IgG N (%) | IgA and IgG N (%) | IgA or IgG<sup>a</sup> N (%) | PR (95% CI) | p-value<sup>b</sup> |
| --- | --- | --- | --- | --- | --- | --- | --- | --- |
| **Gender** |  |  |  |  |  |  |  |  |
| Female | 162 (57.9) | 03-83 | 83 (51.2) | 95 (58.6) | 77 (47.5) | 101 (62.3) | 0.92 (0.77-1.09) | 0.207 |
| Male | 118 (42.1) | 01-79 | 73 (61.9) | 75 (63.6) | 68 (57.6) | 80 (67.8) | 1.09 (0.92-1.29) | 0.207 |
| **Age** |  |  |  |  |  |  |  |  |
| 1-12 | 39 (13.9) |  | 20 (51.3) | 24 (61.5) | 20 (51.3) | 24 (61.5) | 0.94 (0.72-1.23) | 0.399 |
| 13-19 | 36 (12.9) |  | 18 (50.0) | 23 (63.9) | 16 (44.4) | 25 (69.4) | 1.09 (0.86-1.38) | 0.323 |
| 20-29 | 39 (13.9) |  | 23 (59.0) | 24 (61.5) | 20 (51.3) | 27 (69.2) | 1.08 (0.86-1.36) | 0.321 |
| 30-39 | 48 (17.1) |  | 18 (37.5) | 25 (52.1) | 18 (37.5) | 25 (52.1) | 0.77 (0.58-1.03) | **0.033\*** |
| 40-49 | 48 (17.1) |  | 35 (72.9) | 33 (68.8) | 33 (68.8) | 35 (72.9) | 1.16 (0.95-1.41) | 0.125 |
| 50-59 | 36 (12.9) |  | 23 (63.9) | 24 (66.7) | 21 (58.3) | 26 (72.2) | 1.14 (0.91-1.42) | 0.203 |
| ≥60 | 34 (12.2) |  | 19 (55.9) | 17 (50.0) | 17 (50.0) | 19 (55.9) | 0.85 (0.62-1.16) | 0.171 |
| **Marital status** |  |  |  |  |  |  |  |  |
| Single | 89 (45.6) | 14-71 | 51 (57.3) | 54 (60.7) | 44 (49.4) | 61 (68.5) | 1.12 (0.91-1.37) | 0.184 |
| Married or living together | 101 (51.8) | 18-83 | 55 (54.5) | 59 (58.4) | 52 (51.5) | 62 (61.4) | 0.90 (0.73-1.11) | 0.204 |
| Widower | 5 (2.6) | 43-71 | 3 (60.0) | 2 (40.0) | 2 (40.0) | 3 (60.0) | 0.93 (0.45-1.91) | 0.399 |
| **Level of schooling** |  |  |  |  |  |  |  |  |
| Up to middle school | 46 (23.6) | 13-83 | 22 (47.8) | 23 (50.0) | 19 (41.3) | 26 (56.5) | 0.83 (0.63-1.10) | 0.111 |
| middle school completed | 19 (9.7) | 15-73 | 11 (57.9) | 11 (57.9) | 10 (52.6) | 12 (63.2) | 0.97 (0.67-1.39) | 0.475 |
| High school incomplete | 45 (23.1) | 14-64 | 25 (55.6) | 26 (57.8) | 22 (48.9) | 29 (64.4) | 0.99 (0.77-1.26) | 0.473 |
| High school completed | 58 (29.7) | 17-68 | 33 (56.9) | 35 (60.3) | 29 (50.0) | 39 (67.2) | 1.05 (0.84-1.30) | 0.406 |
| In higher education | 27 (13.9) | 18-79 | 19 (70.4) | 21 (77.8) | 19 (70.4) | 21 (77.8) | 1.23 (0.98-1.56) | 0.103 |
| **COVID-19 information sources** |  |  |  |  |  |  |  |  |
| TV | 63 (32.3) | 14-83 | 31 (49.2) | 35 (55.6) | 29 (46.0) | 37 (58.7) | 0.86 (0.68-1.09) | 0.129 |
| Internet | 20 (10.3) | 13-56 | 11 (55.0) | 11 (55.0) | 9 (45.0) | 13 (65.0) | 1.00 (0.71-1.40) | 0.407 |
| TV and Internet | 34 (17.4) | 16-79 | 16 (47.1) | 18 (52.9) | 14 (41.2) | 20 (58.8) | 0.89 (0.65-1.20) | 0.258 |
| Others (religious leader or friends) | 78 (40) | 13-75 | 52 (66.7) | 52 (66.7) | 47 (60.3) | 57 (73.1) | 1.22 (1.00-1.49) | **0.04\*** |
| **Frequency of leaving the home** |  |  |  |  |  |  |  |  |
| Rarely | 34 (17.4) | 14-74 | 16 (47.1) | 17 (50.0) | 15 (44.1) | 18 (52.9) | 0.78 (0.56-1.09) | 0.075 |
| Once a week | 102 (52.3) | 13-83 | 57 (55.9) | 59 (57.8) | 51 (50.0) | 65 (63.7) | 0.96 (0.78-1.17) | 0.390 |
| 2-4 times a week | 59 (30.3) | 15-64 | 37 (62.7) | 40 (67.8) | 33 (55.9) | 44 (74.6) | 1.22 (1.00-1.49) | **0.048\*** |

PR = Prevalence Ratio; CI = Confidence Interval; p<0.05 (\*); <sup>a</sup>Fisher Exact; <sup>b</sup>Yates corrected X<sup>2</sup>

The level of education of the study population was considered low. Only 29.7% of the study population had received a high school diploma (Table 1). Susceptibility to the SARS-CoV-2 infection was greater among the individuals for whom the main source of COVID-19-related information was religious leaders or friends (95% CI: 1.00-1.49; p=0.040). The level of social isolation was directly related to the increased risk of infection. People who declared leaving the home 2-4 times a week were more susceptible to infection (95% CI: 1.00-1.49; p=0.044) (Table 1). The main reasons for leaving the home were either to work or buy food.

As a typical Brazilian indigenous house is small and overcrowded, since usually it serves as a collective house (2 or more families), we evaluated the influence of dwelling size on the prevalence of SARS-CoV-2 infection. Our findings demonstrate that the higher the number of individuals per house the higher the susceptibility to SARS-CoV-2 infection. The susceptibility of people living with 5 people or more is almost 4-fold higher when compared with people living with 2 people and almost 5-fold higher when compared with people living with one other person (95% CI: 1.96-6.01; p=0.010) (Figure 4A). The prevalence observed also progressively increased as the number of individuals per home rose (Figure 4B). The highest prevalence (64.77%) was observed among individuals living with 5 or more people.

**Figure 4.**
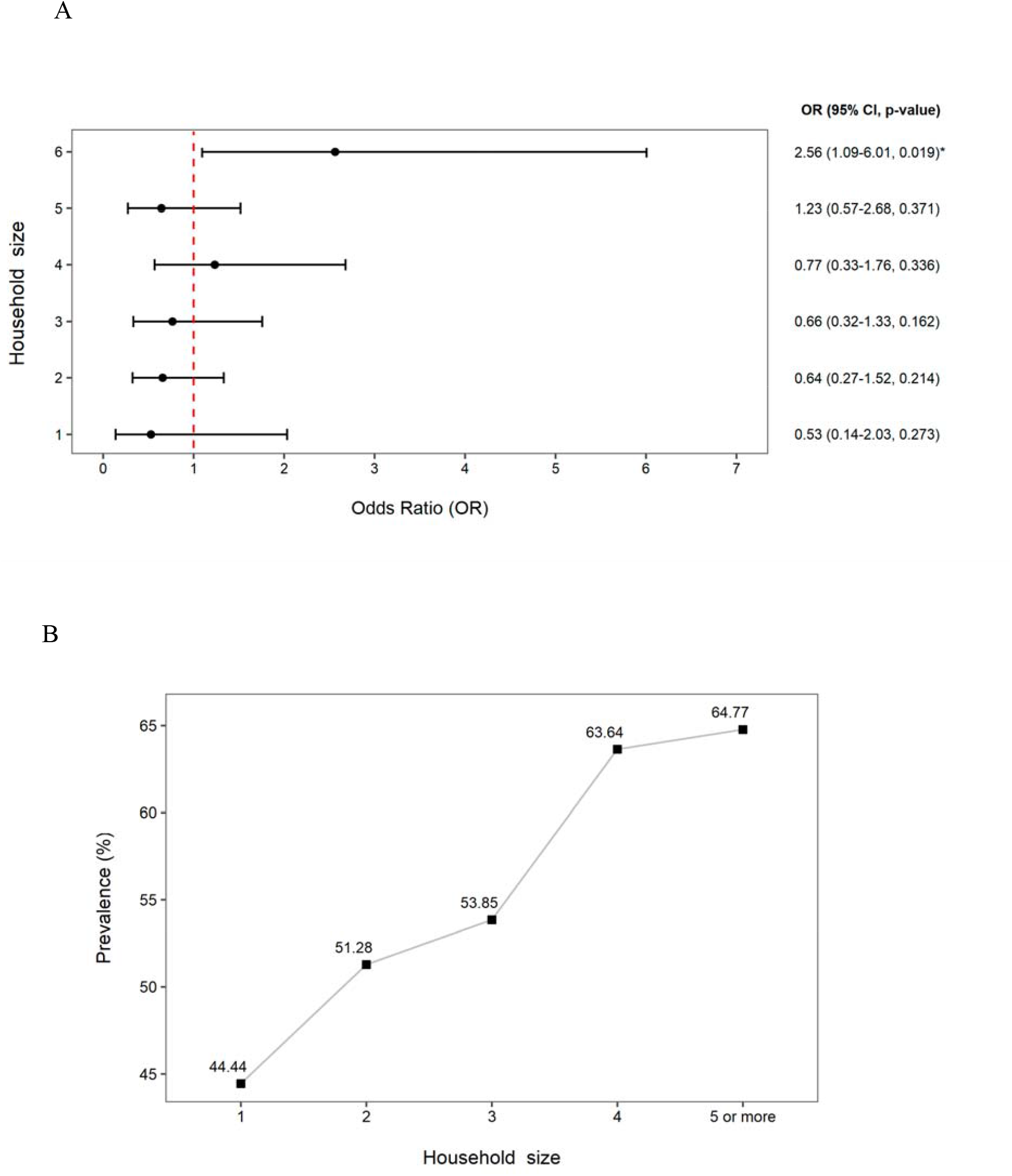
Prevalence rates by household size. (A) Correlation between the risk of exposure to infection and the household size (number of members). (B) SARS-CoV-2 prevalence rates according to the number of inviduals per resindence. OR Fisher’s exact test (* p<0.05)

**Figure 5.**
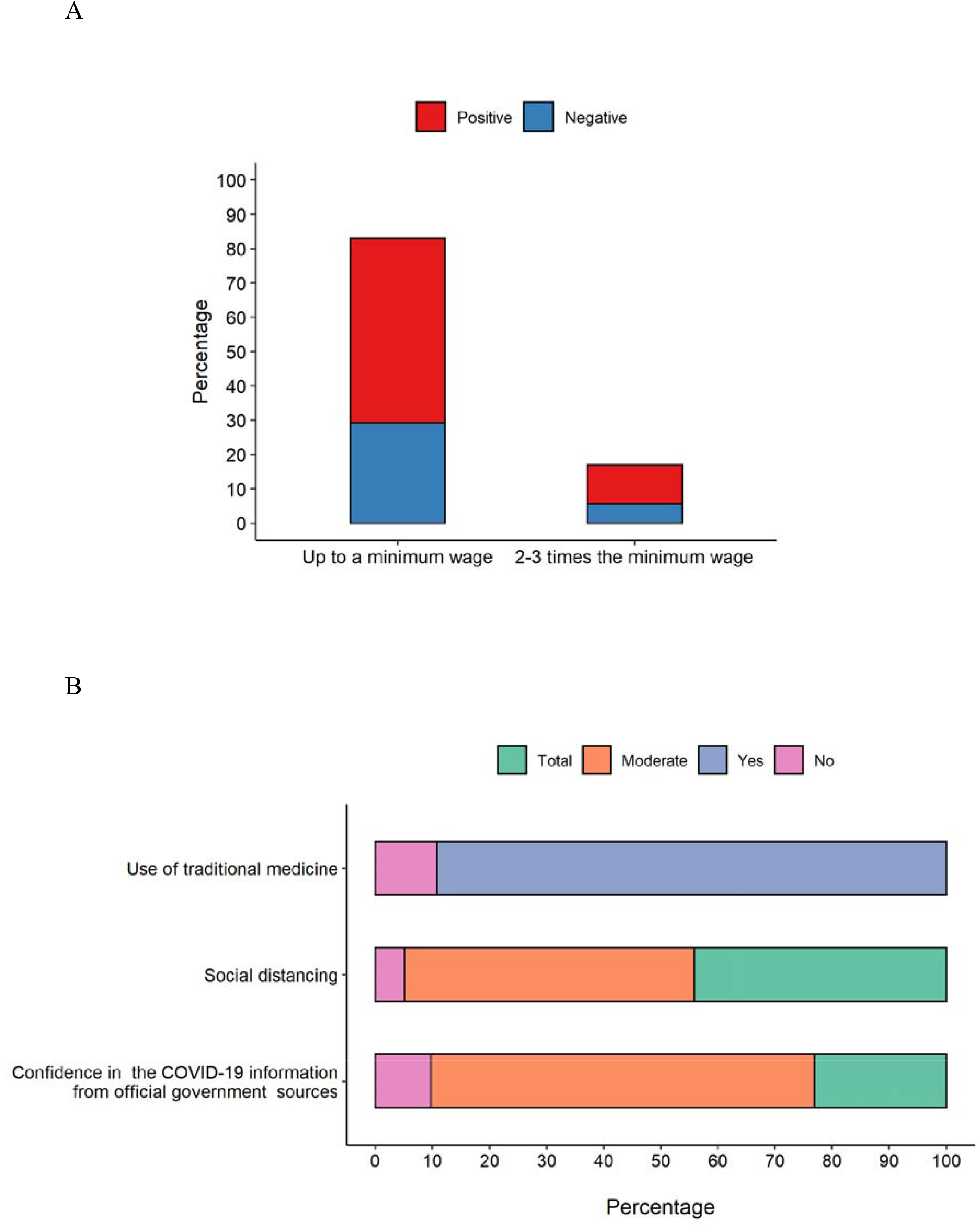
(A) Purchasing power and behavior characteristics (B) of study population.

The level of purchasing power of the study population was very low. Over 83% of individuals earn up to one minimum wage (approximately $192,00 US dollars in Brazil), which hampers effective access to adequate nutrition and healthcare assistance (Figure 4A). Additionally, the majority of the study population (90%) declared using traditional medicine to treat or prevent COVID-19 and other infectious diseases. Most of the individuals (>60%) declared that they partially trust the official Brazilian government COVID-19-related information. According to their answers, only 5.13% did not practice social distancing.

## 4. Discussion

The Brazilian northern region poses the highest rate of SARS-CoV-2 infection vulnerability. By late March 2021, Amazonas state concentrated the most deaths per million people (9). As such, the effectiveness of pandemic response management demands a better comprehension on the impact of social asymmetries in relation to SARS-CoV-2 exposure risk within different population groups, especially amid ethnic minorities living in poverty, such as the indigenous populations of this region.

To our knowledge, this is the first report on the epidemiological situation of SARS-CoV-2 infection in a Brazilian indigenous urban community. Our findings demonstrate that SARS-CoV-2 is highly disseminated among the study population since it showed a prevalence rate of 60.11%. Similar prevalence was observed in the rural indigenous village of Xikrin of Bacajá (Kayapó), in the Brazilian state of Pará, which is also in the northern region (10). Moreover, a population-based study conducted in Manaus during the same period of our study identified a prevalence of 30.34% while the Brazilian National SARS-CoV-2 prevalence estimated in June of 2020 was 3.1% (11,12). These data clearly show that indigenous people are disproportionately hit by SARS-CoV-2 infection. This scenario might be a result of social vulnerabilities associated with cultural practices, which in turn maximize dissemination of the virus and put this population at greater risk.

SARS-CoV-2 strikes ethnic minorities who experience multidimensional poverty much harder, principally the indigenous peoples that constitute about 15% of the population living in extreme poverty in Brazil (13). This population struggles to adopt measures to prevent and mitigate the SARS-CoV-2 spread since they have limited access to basic services like adequate sanitation and clean water. The study population was composed of families of low education and income levels. Our findings showed that over 95% of the residents of Parque das Tribos had no access to clean drinking water and adequate sanitation. The precarious and chaotic social conditions of the study population could be the main reason for the high positivity rates observed.

The positivity rates for anti-SARS-CoV-2 IgA Abs found in this study was 55%. IgA is a dimeric polyvalent antibody found predominantly in mucosal surfaces of humans and other mammals (14). The role of this antibody in the course of SARS-CoV-2 infection is not completely understood, but it might be crucial to the pathogenesis of COVID-19. IgA seems to more effectively neutralize the SARS-CoV-2 than other antibody subclasses (15). Furthermore, serum IgA appears earlier than IgM and IgG in SARs-CoV-2 infected individuals and may persist for several weeks after complete recovery (16). In the present study, a high IgA ratio was detectable in almost 90% of individuals after more than four weeks of the onset of their symptoms. All individuals with symptoms on the day of blood collection were positive for both IgA and IgG. It is possible that the seroconversion for IgA and IgG happened simultaneously in these individuals, as previously reported (17). From the total number of infected individuals, 80.11% were positive for both IgA and IgG. As IgA has a high potential for virus neutralization and may last for a long time, the course of SARS-CoV-2 infection could be influenced by its serum levels. However, we did not follow-up the study population to observe the clinical outcome of the infection. More comprehensive studies are necessary in order to confirm this speculation.

The anti-SARS-CoV-2 IgA and IgG ratios gradually rose with age and elderly individuals (≥ 60 years old) showed the highest antibody ratios. However, the age group with the highest risk of SARS-CoV-2 infection was that of 30-39 years old, just as observed in the non-indigenous Brazilian population (12). Probably these individuals are more susceptible or exposed to SARS-CoV-2 infection because they are working adults who must leave home to support their families. A nationwide study conducted in the United States showed that working adults (20-49 years old) are a higher transmission risk (18). Indeed, the individuals of the study population who left home more often (2-4 times a week) had an increased infection risk. The main reasons for leaving home were either to work or to buy food.

Housing is also an important factor that affects SARS-CoV-2 dissemination among indigenous people. Keeping your distance from others in an indigenous home is not often feasible because they are often overcrowded and very small, which enhances the transmission of the virus (9). Our findings demonstrated that individuals living in households with six or more people had an almost 5-fold increase in risk. This situation is worsened by poor-quality housing. The dwellings of Parque das tribos are very small buildings constructed with low-quality materials, without essential basic infrastructure such as a supply of safe drinking water or effective sewerage. Thus, housing conditions are completely inadequate for keeping them safe from many communicable diseases, including COVID-19.

The majority of the study population declared to adopt distancing measures during the pandemic. However, this might be a huge challenge to these individuals due to their socioeconomic liabilities along with the difficult access to COVID-19-related information (19). Although access to information is a human right, providing accurate information about COVID-19 to indigenous peoples is very complicated. They face difficulties due to language barriers and in accessing different platforms to get updated information. As a consequence, many individuals do not trust the official government information and consult religious leaders for guidance related to different aspects of their lives, including health issues (20). Our data also showed that most of the individuals partially trust the information coming from the Brazilian government and those who get COVID-19-related information from religious leaders or friends are at increased risk for SARS-CoV-2 infection.

Furthermore, about 90% of the study population reported using medicinal plants or herbs to treat or prevent SARS-CoV infection. The use of medicinal herbs is an important part of indigenous culture and must be respected. Traditional knowledge is the foundation of the available conventional treatments and is the cornerstone of pharmacological research (21,22). However, the use of herbs can cause a false sensation of protection, which may result in the discontinuation of important measures to prevent COVID-19 and enhance the vulnerability of these populations.

Despite the limitation of our data, this study indicates that indigenous peoples living in urban areas are being dramatically affected by SARS-CoV-2, especially because of their poor socioeconomic conditions and lack of access to adequate health assistance. This population is fighting a double battle due to the fact that a) in the Brazilian national health system (SUS), their indigenous identity is not recognized by the patient management system and b) They cannot be assisted by SESAI because they are outside their villages or reservations. Both situations reinforce the invisibility of these populations. Thus, we need coordinated national actions that prioritize ethnic vulnerable groups in the battle against COVID-19. We need public policies that promote health, adequate housing and sanitation for these populations. Otherwise, indigenous people living in urban areas are doomed to suffer on unprecedented levels during the current pandemic.

## Data Availability

All data generated or analyzed during this study are included in the main article or its supplementary information files.

## Acknowledgments

The authors thank the community chief Miqueias Kokama and the community residents Vanderlecia Ortega dos Santos, Milton Clézio Gaspar Fidelis, Elizangela Pastor Machado de Freitas, Marigilda da Silva Melgueiro, Cristina Quirino Mariano, Alcinea Martins Albuquerque and Ana Cláudia Martins Tomas for all support provided during this study.

## Authors’ Contributions

Conceptualization: Gemilson Soares Pontes, Ana Carla Bruno, Luciano Cardenes Santos and Antônio de Pádua Quirino Ramalho; methodology: Jean de Melo Silva, Carlos Eduardo de Castro Alves, Leonardo Calheiros de Oliveira and Danielle Furtado da Silva; formal analysis: Gemilson Soares Pontes,Anderson Nogueira Barbosa, Jean de Melo Silva and Renato Pinheiro-Silva; investigation: Antônio de Pádua Quirino Ramalho, Gemilson Soares Pontes, Ana Carla Bruno, Luciano Cardenes Santos, Jean de Melo Silva, Renato Pinheiro-Silva and Danielle Furtado da Silva; validation: Gemilson Soares Pontes and Anderson Nogueira Barbosa; resources: Gemilson Soares Pontes and Allyson Guimarães da Costa; data curation: Gemilson Soares Pontes and Anderson Nogueira Barbosa; writing—original draft preparation: Gemilson Soares Pontes,Jean de Melo Silva and Renato Pinheiro-Silva; writing—review and editing: Gemilson Soares Pontes and Allyson Guimarães da Costa; supervision: Gemilson Soares Pontes; project administration, Gemilson Soares Pontes; funding acquisition: Gemilson Soares Pontes and Allyson Guimarães da Costa. All authors have read and agreed to the published version of the manuscript.

## Figures Legends

**Figure Supplementary 1.**
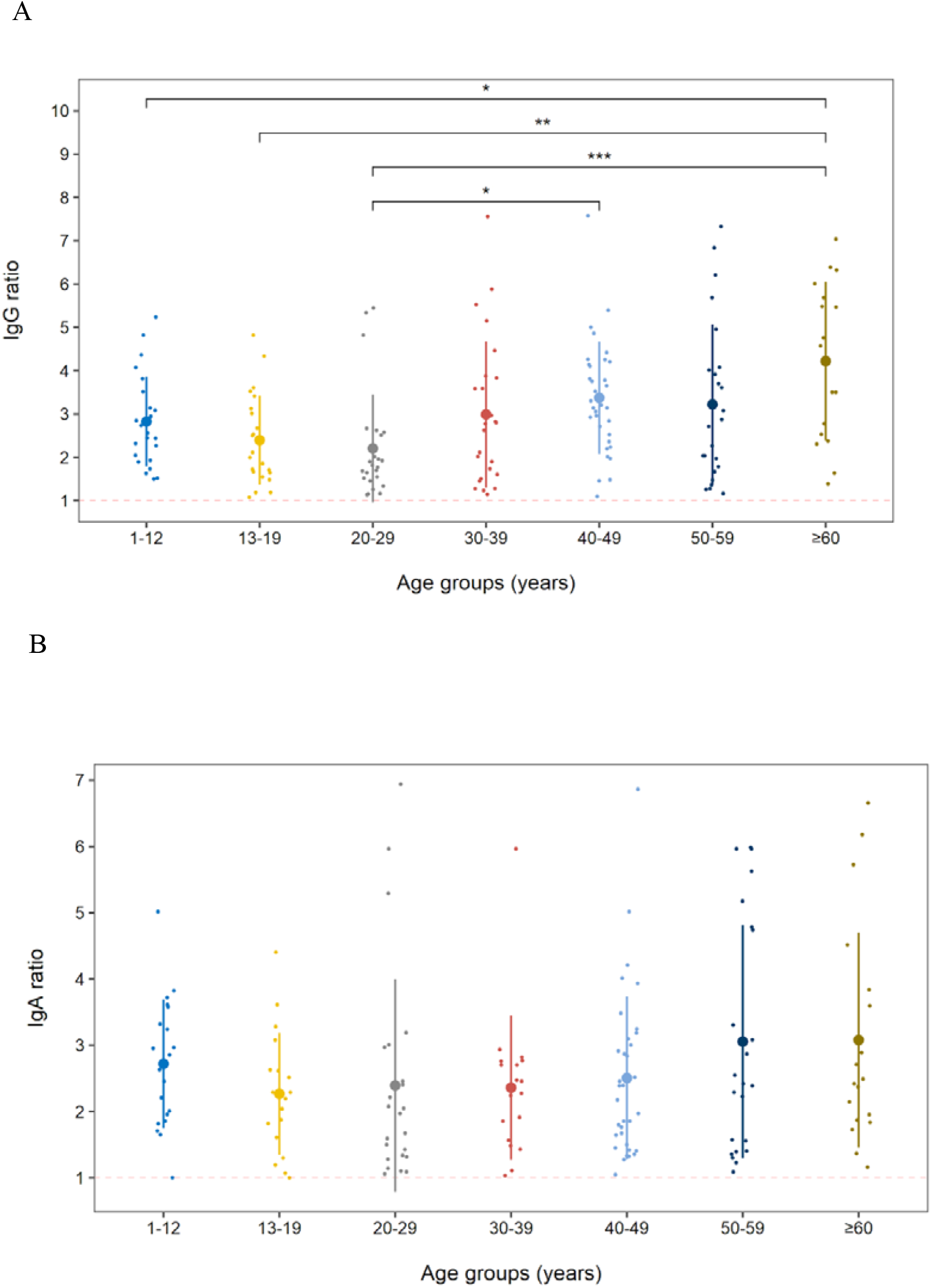
SARS-CoV-2 positivity based on the antibody ratios of age groups. Difference of IgG (A) and IgA (B) ratios between the age groups. Smaller dots represent the antibody ratio from each individual. Larger dots and vertical lines represent mean and standard deviation, respectively. Dashed line represent cut-off values specified by the manufacturer. One-way ANOVA (p < 0.001) with Tukey’s post hoc test (* p<0.05, ** p<0.01, *** p<0.001).

## REFERENCES

1. Amazonas F. Boletim diário COVID-19 no Amazonas 23/3/2021. 2021;354:http://www.fvs.am.gov.br/transparenciacovid19_dado.

2. Moran EF, Brondízio ES, VanWey LK. Population and environment in Amazônia: Landscape and household dynamics. In: Population, Land Use, and Environment: Research Directions. 2005. p. 5.

3. Instituto Brasileiro de Geografia e Estatistica (IBGE). População estimada. Diretoria de Pesquisas, Coordenação de População e Indicadores Sociais, Estimativas da população residente com data de referência 1o de julho de 2019. 2020.

4. IBGE. Senso demográfico IBGE. Instituto Brasileiro de Geografia e Geoestatistica. 2010.

5. Meneses-Navarro S, Freyermuth-Enciso MG, Pelcastre-Villafuerte BE, Campos-Navarro R, Meléndez-Navarro DM, Gómez-Flores-Ramos L. The challenges facing indigenous communities in Latin America as they confront the COVID-19 pandemic. International Journal for Equity in Health. 2020.

6. Saúde M da, Indígena. SE de S. Boletim epidemiológico. 2021.

7. Pereira JCM. Indígenas na cidade de Manaus (AM). Novos Cad NAEA. 2021;23(3):11–31.

8. Borgers N, Hox J, Sikkel D. Response quality in survey research with children and adolescents: The effect of labeled response options and vague quantifiers. Int J Public Opin Res. 2003;15(1):83–94.

9. Tavares FF, Betti G. The pandemic of poverty, vulnerability, and COVID-19: Evidence from a fuzzy multidimensional analysis of deprivations in Brazil. World Dev. 2021;139.

10. Rodrigues EPS, Abreu IN, Lima CNC, da Fonseca DLM, Pereira SFG, dos Reis LC, et al. High prevalence of anti-SARS-CoV-2 IgG antibody in the Xikrin of Bacajá (Kayapó) indigenous population in the brazilian Amazon. International Journal for Equity in Health. 2021.

11. Lalwani P et al. SARS-CoV-2 seroprevalence and associated factors in Manaus, Brazil:baseline results from the DETECTCoV-19 cohort study. 2021;

12. Hallal PC, Hartwig FP, Horta BL, Silveira MF, Struchiner CJ, Vidaletti LP, et al. SARS-CoV-2 antibody prevalence in Brazil: results from two successive nationwide serological household surveys. Lancet Glob Heal. 2020;8(11):e1390–8.

13. Nathaniel Berger D, Bulanin N, García-Alix L, Wiben Jensen M, Leth S, Alvarado Madsen E, et al. 2 IWGIA-The Indigenous World-2020. In.

14. Woof JM, Russell MW. Structure and function relationships in IgA. Mucosal Immunol. 2011;(4):590–597.

15. Wang Z, Lorenzi JCC, Muecksch F, Finkin S, Viant C, Gaebler C, et al. Enhanced SARS-CoV-2 neutralization by dimeric IgA. Sci Transl Med. 2021;3(577):eabf1555.

16. Yu HQ, Sun BQ, Fang ZF, Zhao JC, Liu XY, Li YM, et al. Distinct features of SARS-CoV-2-specific IgA response in COVID-19 patients. Eur Respir J. 2020;56(2):2001526.

17. Long QX, Liu BZ, Deng HJ, Wu GC, Deng K, Chen YK, et al. Antibody responses to SARS-CoV-2 in patients with COVID-19. Nat Med. 2020;(26):845–8.

18. Monod M, Blenkinsop A, Xi X, Hebert D, Bershan S, Tietze S, et al. Age groups that sustain resurging COVID-19 epidemics in the United States. Science (80-). 2021;1336(March):eabe8372.

19. Cupertino GA, do Carmo Cupertino M, Gomes AP, Braga LM, Siqueira-Batista R. COVID-19 and Brazilian indigenous populations. Am J Trop Med Hyg. 2020;103(2):609–12.

20. Oliveira RNDC, Rosa LCDS. SÁUDE INDÍGENA EM TEMPOS DE BARBÁRIE: política pública, cenários e perspectivas. Rev Políticas Públicas. 2015;18(2):481.

21. Sharma M, Anderson SA, Schoop R, Hudson JB. Induction of multiple pro-inflammatory cytokines by respiratory viruses and reversal by standardized Echinacea, a potent antiviral herbal extract. Antiviral Res. 2009;83(2):165–70.

22. Hudson J, Vimalanathan S. Echinacea-A source of potent antivirals for respiratory virus infections. Pharmaceuticals. 2011;4(7):1019–31.

